# Extubation generates lung volume inhomogeneity in preterm infants

**DOI:** 10.1101/2021.02.03.21251050

**Authors:** R Bhatia, HR Carlisle, RK Armstrong, COF Kamlin, PG Davis, DG Tingay

**Author notes:** **Corresponding Author:** Dr. Risha Bhatia, Monash Newborn, Monash Children’s Hospital, 246, Clayton Road, Clayton, VIC 3168.

## Abstract

**Objective:** To evaluate the feasibility of EIT to describe the regional tidal ventilation (V_T_) and change in end-expiratory lung volume (EELV) patterns in preterm infants during the process of extubation from invasive to non-invasive respiratory support.

**Design:** Prospective observational study

**Setting:** Single-centre tertiary neonatal intensive care unit

**Patients:** Preterm infants born <32 weeks gestation who were being extubated to nasal continuous positive airway pressure (nCPAP) as per clinician discretion.

**Interventions:** Electrical Impedance Tomography measurements were taken in supine infants during elective extubation from synchronised positive pressure ventilation (SIPPV) before extubation, during and then at 2 and 20 minutes after commencing nCPAP. Extubation and pressure settings were determined by clinicians.

**Main outcome measures:** Global and regional ΔEELV and ΔV_T_ were measured. Heart rate, respiratory rate and oxygen saturation were measured throughout.

**Results:** Thirty infants of median (range) 2 (1, 21) days were extubated to a median (range) CPAP 7 (6, 8) cmH_2_O. SpO_2_/FiO_2_ ratio was mean (95% CI) 50 (35, 65) lower 20 minutes after nCPAP compared with SIPPV. EELV was lower at all points after extubation compared to SIPPV, and EELV loss was primarily in the ventral lung (p=0.04). V_T_ was increased immediately after extubation, especially in the central and ventral regions of the lung, but the application of nCPAP returned V_T_ to pre-extubation patterns.

**Conclusions:** Lung behaviour during the transition from invasive positive pressure ventilation to CPAP at moderate distending pressures is variable and associated with lung volume loss in the ventral lung.

## Introduction

Minimising exposure to invasive respiratory support via an endotracheal tube (ETT) reduces the risk of lung injury in preterm infants.^1^ Despite attempts to avoid intubation, approximately 50% infants born <32 weeks’ gestation still receive invasive respiratory support.^2^ Nasal continuous positive applied pressure (nCPAP) and caffeine therapy allows earlier extubation.^3,4^ Even with the use of nCPAP, 40% of preterm infants experience extubation failure.^2,5^ This has led to a focus on identifying factors that may predict successful extubation. However, beyond broad criteria such as gestation, weight and oxygen need, predictive tools have had limited impact.^5,6^

The causes of extubation failure in preterm infants are multifactorial,^5^ but atelectasis and impaired gas exchange are often the final common manifestations. Impaired central respiratory drive, fatigue, residual primary lung disease and secondary events such as sepsis are common associations. The physiology of the transition from invasive respiratory support via an ETT to non-invasive ventilation may provide insights on the underlying mechanisms of extubation failure but is poorly understood, partly because imaging of this process has been very difficult. Recent development of non-invasive, bedside tools, specifically electrical impedance tomography (EIT) and lung ultrasound, has allowed closer examination of the changes occurring around the time of extubation.^7,8^ These have shown complex regional ventilation and aeration patterns occurring even in preterm infants considered stable, with relatively low fraction of inspired oxygen (FiO_2_), on nCPAP.^9^

This study aimed to evaluate the feasibility of EIT to describe the regional tidal ventilation (V_T_) and end-expiratory lung volume (EELV) patterns in preterm infants during the process of successful elective extubation to nCPAP. To address these aims, we analysed our dataset of EIT measurements from preterm infants made as part of a series of studies conducted between 2008-2013.^10-12^

## Methods

This prospective observational study was performed in the Neonatal Intensive Care Unit (NICU) of the Royal Women’s Hospital, Melbourne, Australia. Prospective written informed parental consent was provided in infants born <32 weeks’ gestation participating in 1) a larger study of regional ventilation during synchronised mechanical ventilation with volume targeting (SIPPV),^10,11^ or 2) studies undertaken as part of a PhD thesis examining regional volume characteristics during nCPAP.^12^ Parental consent was reaffirmed prior to extubation for infants participating in the first study. These infants were included if an investigator was available to record EIT measurements during elective extubation from SIPPV to short bi-nasal prong nCPAP before 28 days of age, and the infant could be nursed supine during extubation. The decision to extubate was made on clinical grounds by the treating team. All infants received caffeine citrate and passed a 3-minute spontaneous breathing test prior to extubation.^6^ Infants were not sedated.

All EIT measurements were taken with the infant in the supine position. Three, 2-minute EIT recordings (GeoMF II EIT system, Cardinal Health, Hoechberg, Germany) of relative impedance change (ΔZ) were made 20-30 minutes prior to anticipated extubation using our previously detailed methodology.^10^ Thereafter, nursing staff prepared the infant for extubation (including fitting the hat and nCPAP circuit onto the infant’s head). Continuous EIT recordings were made from immediately prior to extubation until 5 minutes after nCPAP had commenced (defined as the nCPAP prongs being appropriately fitted in the nares). Following extubation there was a variable period before the nCPAP prongs were placed in the nares during which the infant received no respiratory support whilst the nursing staff fitted the nasal prongs and performed other immediate post-extubation tasks, such as oral suction. During this time, nursing staff could provide T-piece mask CPAP support at their discretion. A final series of three, 2-minute EIT recordings were made between 15 and 25 minutes after the infant was established on nCPAP. Delivered FiO_2_ and relevant ventilator parameters were recorded from the ventilator (Dräger BabyLog 8000*Plus*, Drägerwerk AG, Lübeck, Germany). Heart rate (HR), peripheral oxygen saturation (SpO_2_) and respiratory rate (RR) were recorded from the bedside MP70 monitor (Phillips, Eindhoven, Netherlands) at 12 second intervals. FiO_2_ was titrated to target SpO_2_ of 88-92% throughout.

EIT and associated cardiorespiratory parameters were analysed during four phases of the extubation process:

1. SIPPV: Quiet breathing 20 minutes prior to extubation
2. No CPAP: The period immediately upon removal of the ETT until the nCPAP prongs were appropriately fitted in the nares
3. CPAP_2min_: 2 minutes after commencing nCPAP (prongs in nares)
4. CPAP_20min_: 20 minutes after commencing nCPAP (prongs in nares)

During SIPPV, CPAP_2min_ and CPAP_20min_ a minimum of 60 inflations/spontaneous breaths of artefact-free continuous EIT data closest to each time point were identified and analysed. All available spontaneous breaths were analysed during the No CPAP phase. Using our previously detailed methods, and in accordance with international standards, EIT data were reconstructed using an appropriate chest model.^7,9^ The reconstructed uncalibrated ΔZ signals were low-pass filtered to the second harmonic of the respiratory rate and EELV and tidal volume (V_T_) calculated from the trough and amplitude of the tidal ΔZ signal.^7,10^ EELV was referenced to the mean EELV impedance value during the SIPPV phase to determine the relative change in EELV (ΔEELV) following extubation. Relative ΔEELVs were calculated for the whole chest, ventral (non-gravity dependent) and dorsal (gravity dependent) halves of the chest. Regional V_T_ was calculated for each of 32 equidistant lung regions from the most ventral (0%) to most dorsal (100%) to create a functional map of gravity-dependent regional V_T_ change in the chest. The gravity-dependent centre of ventilation was calculated at each phase.^7,9,13,14^ All regional contributions were adjusted to the anatomical size of each region.^7^

Descriptive statistics were generated for regional and global EIT data and comparisons made using repeated measure one-way ANOVA or paired t-test. Subtraction histograms of the difference between V_T_ within each gravity-dependent region for No CPAP, CPAP_2min_ and CPAP_20min_ against SIPPV were generated.^15^ Statistical analysis was performed using Prism (V9.0, GraphPad, CA, USA) and a p value <0.05 considered significant.

## Results

Thirty infants were studied at extubation and had complete datasets for analysis (see Online Supplementary Figure for the study flow chart). Their characteristics are summarised in Table 1. No infant required intubation within 24 hours. There was no difference in heart rate and respiratory rate at each phase. The SpO_2_/FiO_2_ ratio was a mean (95% CI) 50 (35, 65) lower at CPAP_20min_ compared with SIPPV (paired t-test), and 21 infants remained in ≤0.30 FiO_2_ throughout. The median (range) length of the No CPAP phase was 61 (8, 285) s.

**Table 1.** Infant characteristics.

|  |  |
| --- | --- |
| n=30 |  |
| Gestational Age | 27.9 (2.0) weeks |
| Age | 2 (1, 21) days |
| Birthweight | 1085 (375) g |
| Weight at extubation | 1085 (376) g |
| Female | 13 (43%) |
| Antenatal Steroids | 30 (100%) |
| Pre-extubation |  |
| PEEP | 5 (5, 6) cmH<sub>2</sub>O |
| VTV | 3.8 (0.5) ml/kg |
| FiO<sub>2</sub> | 0.22 (0.21, 0.40) |
| SpO<sub>2</sub>/FiO<sub>2</sub> | 400 (73) |
| pH | 7.33 (0.06) |
| PcCO<sub>2</sub> | 46.9 (9.7) mmHg |
| Heart Rate | 154 (12) bpm |
| Respiratory rate | 65 (14) bpm |
| Surfactant | 27 (90%) |
| Caffeine | 29 (97%) |
| Treated PDA | 6 (20%) |
| 20-min post extubation |  |
| CPAP | 7.0 (6, 8) cmH<sub>2</sub>O |
| FiO<sub>2</sub> | 0.25 (0.21, 0.55) |
| SpO<sub>2</sub>/FiO<sub>2</sub> | 356 (95)* |
| Heart Rate | 155 (12) bpm |
| Respiratory rate | 57 (17) bpm |

All data mean (SD) except; age, PEEP, CPAP and FiO<sub>2</sub> median (range). \* p=0.0006 paired t test (pre-extubation)
SpO<sub>2</sub>/FiO<sub>2</sub>; modified oxygenation index, PcCO<sub>2</sub>; partial pressure of capillary CO<sub>2</sub> at most recent blood gas analysis, PDA; patent ductus arteriosus, PEEP; positive end-expiratory pressure, VTV; volume targeted ventilation, bpm; breath or beat per minute.

Following extubation global EELV decreased but was highly variable, with the lowest EELV at CPAP_20min_ (Figure 1); p=0.08 (repeated measure one-way ANOVA). Extubation was associated with gravity-dependent heterogeneity of EELV. There was very little change in EELV from pre-extubation SIPPV values within the dorsal lung (p=0.67). In contrast, ventral lung ΔEELV decreased significantly after extubation (p=0.04), and accounted for the majority of the global EELV changes.

**Figure 1.**
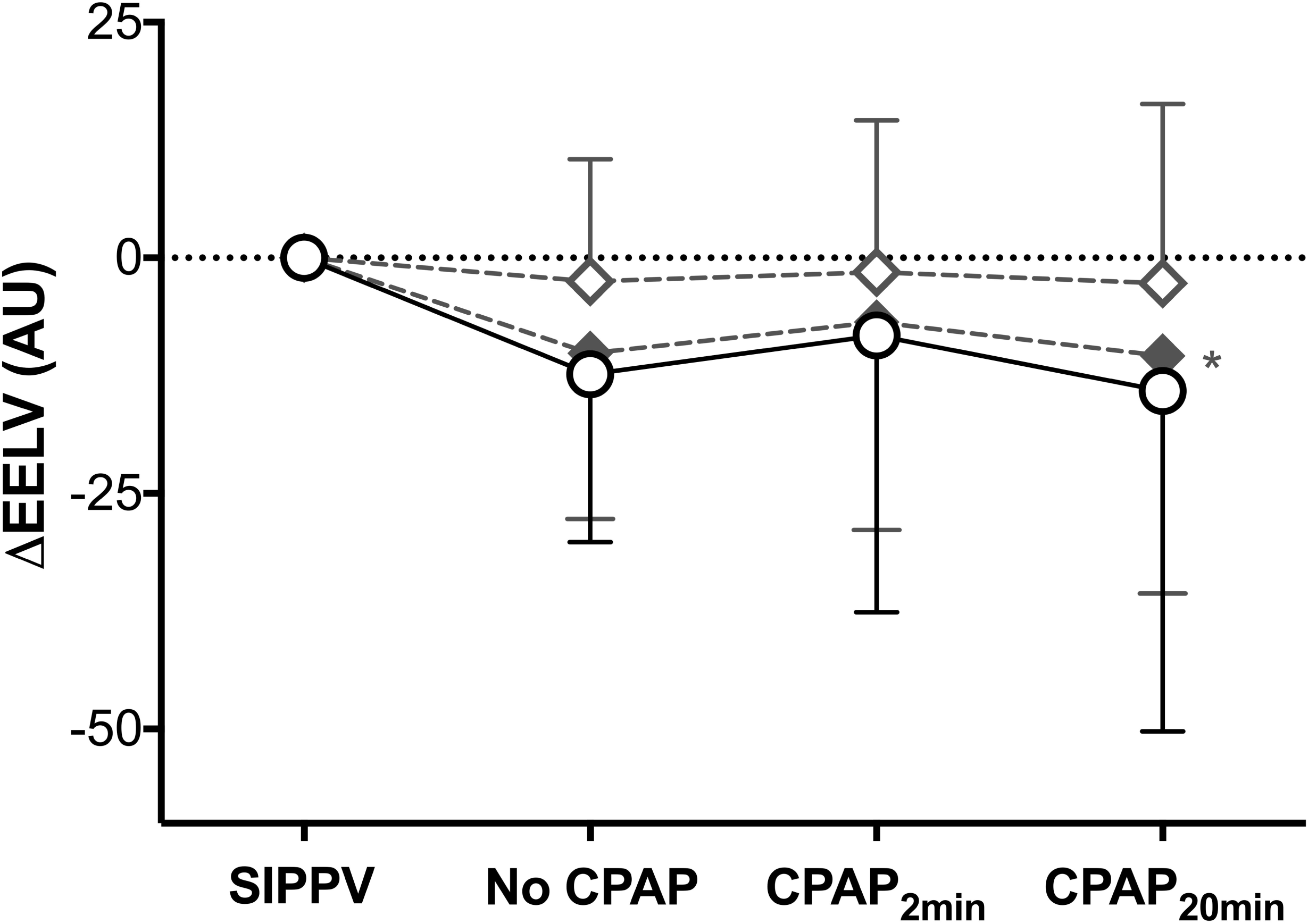
Relative change in end-expiratory lung volume (ΔEELV) from values during SIPPV (arbitrary units; AU) immediately after extubation for the whole chest (black open circles), ventral (closed grey diamonds) and dorsal (open grey diamonds) halves of the chest. ΔEELV after removal of ETT but prior to nasal CPAP commenced (No CPAP), 2 min (CPAP_2min_) and 20 min (CPAP_20min_) after nasal CPAP was commenced. All data mean ± SD. * p=0.040 (ventral); one-way repeated measure ANOVA.

Figure 2 shows the gravity-dependent distribution of V_T_. Extubation was associated with an increase in V_T_ during the No CPAP phase, mostly within the central and ventral regions. The distribution of V_T_ approximated SIPPV patterns at CPAP_2min_ and CPAP_20min_, with slight increases in dorsal lung V_T_. Overall, there was no difference in the gravity-dependent centre of ventilation at each phase; mean (SD) SIPPV 52.6 (1.4)%, No CPAP 52.8 (0.1)%, CPAP_2min_ 52.3 (1.5)% and CPAP_20min_ 52.2 (1.3)%, p=0.62 (repeated measure one-way ANOVA).

**Figure 2.**
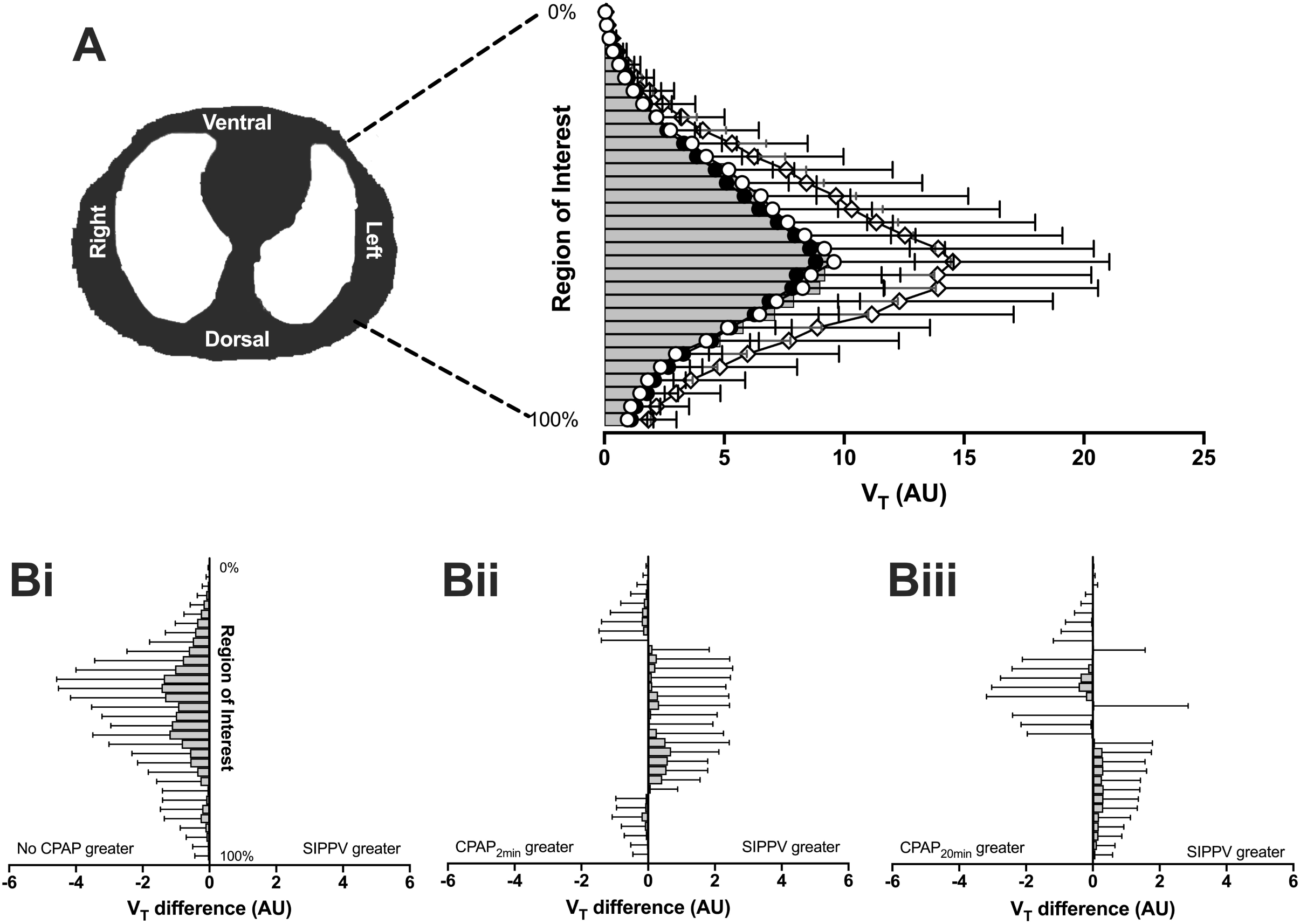
A. Distribution of tidal volume (V_T_) within 32 equally sized regions of the chest along the gravity-dependent plane during SIPPV (grey bars), No CPAP (open diamonds), CPAP_2min_ (open circles) and CPAP_20min_ (black circles). 0% represents the most ventral region and 100% the most dorsal region as illustrated in the insert schematic of the lung regions of chest. **B**. Difference in V_T_ within each lung region between SIPPV and No CPAP (**i**), CPAP_2min_ (**ii**) and CPAP_20min_ (**iii**). All data mean ± SD.

## Discussion

In our observational study of preterm infants, EIT was able to identify changes in regional volume states during successful extubation to nCPAP. A loss of EELV was common, persisted until 20 min after nCPAP had begun, and was associated with gravity dependent heterogeneity of aeration (EELV) but not V_T_. The only previous study of EELV pre and post-extubation in preterm infants compared nCPAP to pre-extubation ETT CPAP without positive pressure inflations, and found an increase in EELV 10 min after starting nCPAP.^16^ The pre-extubation management explains the differences in ΔEELV with our study. During SIPPV, EELV is assisted with positive inflating pressures during active inspiration (and PEEP during passive expiration).^13^ Thus, increasing intrathoracic pressure compared with ETT CPAP in which tidal ventilation is generated solely by negative inflating pressures. In addition, during ETT CPAP the glottis is bypassed, removing the important role it plays in maintaining intra-thoracic volume during expiration (intrinsic PEEP).^17^ Interestingly, EELV remained below pre-extubation values after 20 min of nCPAP, presumably due to the innate differences in transmissible pressures between invasive and non-invasive support. Measurements beyond 20 minutes would be useful to determine whether nCPAP eventually regains global EELV.

The pattern of EELV within the lung changed following extubation. Overall, the ventral lung contributed to 74% of EELV loss at CPAP_20min_. Preterm infants predominantly use their diaphragm to generate tidal ventilation, and previous EIT studies in infants have shown that the dorsal lung is preferentially ventilated during spontaneous breathing.^9,13,14^ The ventral-dorsal shape of the diaphragm is elliptical. Thus, there is greater movement of the dorsal portion of the diaphragm.^18^ In addition there is a greater lung mass in the dorsal thorax. In contrast, positive pressure ventilation via an ETT creates pressure and gas flow conditions that favour the ventral lung and negate some of the greater dorsal diaphragmatic contribution to ventilation.^13^ As such, the dorsal-dominant inhomogeneity is not surprising. However, oxygenation was lower following extubation which suggests functional atelectasis was present and contributed in part to the ΔEELV and inhomogeneity. The role of the diaphragm in preterm respiratory function is often overlooked clinically.^18,19^ We hypothesise that the engagement of dorsal aeration may be an important component of preventing significant atelectasis following extubation. It will be interesting to see if the recently available modes of non-invasive support using diaphragmatic triggering contribute to post extubation success.

Unlike EELV, the relative magnitude and regional distribution of V_T_ were not different during nCPAP and invasive support. nCPAP reduces work of breathing and stabilises airways,^20,21^ effects that were likely present in our study given all infants were stable on nCPAP in the 24 hours after extubation. Our study was limited to a cumbersome research EIT unit.^7^ Despite this, we could demonstrate that EIT may be useful in monitoring the lung during extubation using the temporal trend of relatively simple validated EIT parameters to define patterns of EELV and tidal ventilation.^7^ Clinical EIT systems with electrode belts^22^ and user interfaces which are simpler to use are now available and can provide continuous monitoring over prolonged periods. This should allow larger, properly powered validation studies of the potential of EIT to guide nCPAP support, and potentially aid in the prediction of extubation success and prevention of extubation failure.

The magnitude and distribution of both regional EELV and V_T_ were highly variable within and between infants at all phases of the study. Breathing is a dynamic process, involving the interaction between changing disease state, lung parenchyma, mechanics, drive and therapies. It is likely some infants adapted to extubation better than others and that for most, the reason for extubation success or failure involves an interplay between all of these factors.^5^ Unfortunately our use of a feasibility sample generated from analysis of available data pooled in previous observational EIT studies precludes interpretation of whether the observed variability represents specific factors. Even though our study was relatively large compared with other non-invasive imaging studies in preterm infants,^7-10,14,16,23,24^ to do so would require a much larger population. Despite this, there is an increasing body of work from bedside imaging tools such as lung ultrasound,^25^ EIT,^9,13,26^ and multiple breath washout tests^27^ demonstrating that clinicians are unaware of the dynamic complexity of breathing with current clinical measures being limited to the extreme events such as apnoea, desaturation, and bradycardia. In our study all infants were successfully extubated. Having a comparative group of infants who required reintubation would provide more insight into lung dynamics.

A unique aspect of this study was the recording of lung volumes immediately after extubation, and before nCPAP support had commenced. Relative V_T_ was greater during this phase than any other, especially in the central-ventral regions of the lung. This indicates greater work of breathing, presumably due to the immediate stress of extubation and handling, and to defend lung volume following the loss of any pressure support and/or poor glottic function secondary to intubation. The study design precludes any interpretation of whether the duration without nCPAP, or use of transient mask CPAP, may influence ventilation stability. The application of a face mask to a very preterm infant whilst being prepared for nCPAP may be suboptimal with variable leak, which may compromise effective pressure transmission to the lungs. To our knowledge, no extubation guidelines focus on how to manage the period between extubation and commencing nCPAP.

Our study design, using post-hoc analysis, cannot address two important clinical questions: 1) when to extubate, and 2) how to optimise post-extubation nCPAP settings; neither of which were standardised. All infants were managed supine to standardise measures however, prone positioning is widely used in NICU, and as we have shown in more recent studies, may improve ventilation patterns.^9,14^

In conclusion, EIT was able to describe the complex lung conditions occurring during extubation to nCPAP, specifically lung volume loss and greater use of the dorsal lung. EIT lung imaging may be useful to guide peri-extubation respiratory support and to potentially reduce the burden of extubation failure in preterm infants.

## Data Availability

De-identified individual participant data, study protocols and statistical analysis codes are available from three months to 23 years following article publication to researchers who provide a methodologically sound proposal, with approval by an independent review committee (learned intermediary). Proposals should be directed to to gain access. Data requestors will need to sign a data access or material transfer agreement approved by MCRI.

## Acknowledgements

The authors wish to thank Associate Professor Susan Donath for statistical advice and Brenda Argus for her assistance with this study.

## Ethics approval

This study was approved by the Royal Women’s Hospital Human Research Ethics Committee, in accordance with the National Health and Medical Research Council Statement of Ethical Conduct in Human Research (2007).

## Author contributions

RB, HRC, RKA, DGT and PGD developed the concept and experimental design. RB, HRC, RKA and DGT were involved in data acquisition, analysis and interpretation. DGT, PGD and COFK supervised the study. RB and DGT drafted the first manuscript and all authors contributed to editing.

## Data sharing

De-identified individual participant data, study protocols and statistical analysis codes are available from three months to 23 years following article publication to researchers who provide a methodologically sound proposal, with approval by an independent review committee (“learned intermediary”). Proposals should be directed to to gain access. Data requestors will need to sign a data access or material transfer agreement approved by MCRI.

## What is already known on this topic

- 40% of preterm infants experience extubation failure despite CPAP and caffeine therapy.
- The causes of extubation failure in preterm infants are multifactorial and clinicians have few methods of delineating these at the bedside.
- Electrical Impedance Tomography (EIT) is a bedside imaging method that offers the potential to provide insights on the underlying mechanisms during extubation success and failure.

## What this study adds

- EIT was able to image the complex lung aeration and ventilation changes occurring during extubation to nCPAP.
- In stable infants, lung volume loss was common, and predominantly in the ventral lung, after extubation and persisted until at least 20 minutes.
- Ventilation homogeneity was maintained following extubation, suggesting increased diaphragmatic function is important during adaptation to non-invasive support.

## References

1. Thebaud B, Goss KN, Laughon M, et al. Bronchopulmonary dysplasia. Nat Rev Dis Primers 2019;5:78.

2. Chow SSW, Le Marsney R, Creighton P, Kander V, Haslam R, Lui K. Report of the Australian and New Zealand Neonatal Network 2015. Sydney ANZNN 2017

3. Higgins RD, Richter SE, Davis JM. Nasal continuous positive airway pressure facilitates extubation of very low birth weight neonates. Pediatrics 1991;88:999–1003.

4. Annibale DJ, Hulsey TC, Engstrom PC, et al. Randomized, controlled trial of nasopharyngeal continuous positive airway pressure in the extubation of very low birth weight infants. J Pediatr 1994;124:455–60.

5. Dargaville PA, Gerber A, Johansson S, et al. Incidence and Outcome of CPAP Failure in Preterm Infants. Pediatrics 2016;138:e20153985.

6. Kamlin CO, Davis PG, Morley CJ. Predicting successful extubation of very low birthweight infants. Arch Dis Child Fetal Neonatal Ed 2006;91:F180–3.

7. Frerichs I, Amato MB, van Kaam AH, et al. Chest electrical impedance tomography examination, data analysis, terminology, clinical use and recommendations: consensus statement of the TRanslational EIT developmeNt stuDy group. Thorax 2017;72:83–93.

8. Raimondi F, Migliaro F, Sodano A, et al. Use of neonatal chest ultrasound to predict noninvasive ventilation failure. Pediatrics 2014;134:e1089–94.

9. Thomson J, Ruegger CM, Perkins EJ, et al. Regional ventilation characteristics during non-invasive respiratory support in preterm infants. Arch Dis Child Fetal Neonatal Ed 2020 doi: 10.1136/archdischild-2020-320449 [published Online First: 2020/11/29]

10. Armstrong RK, Carlisle HR, Davis PG, et al. Distribution of tidal ventilation during volume-targeted ventilation is variable and influenced by age in the preterm lung. Intensive care medicine 2011;37:839–46.

11. Carlisle HR, Armstrong RK, Davis PG, et al. Regional distribution of blood volume within the preterm infant thorax during synchronised mechanical ventilation. Intensive Care Med 2010;36:2101–8.

12. Bhatia R. Optimising Continuous Positive Airway Pressure in Preterm Infants. University of Melbourne, 2015.

13. Dowse G, Perkins E, Thomson J, et al. Synchronized Inflations Generate Greater Gravity-Dependent Lung Ventilation in Neonates. J Pediatr 2021;228:24–30 e10.

14. Schinckel NF, Hickey L, Perkins EJ, et al. Skin-to-skin care alters regional ventilation in stable neonates. Arch Dis Child Fetal Neonatal Ed 2021;106:76–80.

15. Tingay DG, Rajapaksa A, Zonneveld CE, et al. Spatiotemporal Aeration and Lung Injury Patterns Are Influenced by the First Inflation Strategy at Birth. Am J Respir Cell Mol Biol 2016;54:263–72.

16. van der Burg PS, Miedema M, de Jongh FH, et al. Changes in lung volume and ventilation following transition from invasive to noninvasive respiratory support and prone positioning in preterm infants. Pediatr Res 2015;77:484–8.

17. Tingay D, Farrell O, Thomson J, et al. Unravelling the complexities of the first breaths of life medRxiv 2020;MEDRXIV/2020/161166 doi: 10.1101/2020.07.29.20161166 [Accepted Am J Resp Crit Care Med 18 Jan 2021]

18. Devlieger H, Daniels H, Marchal G, et al. The diaphragm of the newborn infant: anatomical and ultrasonographic studies. J Dev Physiol 1991;16:321–9.

19. Liang F, Emeriaud G, Rassier DE, et al. Mechanical ventilation causes diaphragm dysfunction in newborn lambs. Crit Care 2019;23:123.

20. Pillow J. Which continuous positive airway pressure system is best for the preterm infant with respiratory distress syndrome? Clin Perinatol 2012;39:483–96.

21. Liptsen E, Aghai ZH, Pyon KH, et al. Work of breathing during nasal continuous positive airway pressure in preterm infants: a comparison of bubble vs variable-flow devices. J Perinatol 2005;25:453–8.

22. Sophocleous L, Frerichs I, Miedema M, et al. Clinical performance of a novel textile interface for neonatal chest electrical impedance tomography. Physiol Meas 2018;39:044004.

23. Miedema M, van der Burg PS, Beuger S, et al. Effect of nasal continuous and biphasic positive airway pressure on lung volume in preterm infants. J Pediatr 2013;162:691–7.

24. Miedema M, de Jongh FH, Frerichs I, et al. Changes in lung volume and ventilation during lung recruitment in high-frequency ventilated preterm infants with respiratory distress syndrome. J Pediatr 2011;159:199–205 e2.

25. Loi B, Vigo G, Baraldi E, et al. Lung Ultrasound to Monitor Extremely Preterm Infants and Predict BPD: Multicenter Longitudinal Cohort Study. Am J Respir Crit Care Med 2020 doi: 10.1164/rccm.202008-3131OC [published Online First: 2020/12/23]

26. Gaertner VD, Waldmann AD, Davis PG, et al. Transmission of Oscillatory Volumes into the Preterm Lung during Noninvasive High-Frequency Ventilation. Am J Respir Crit Care Med 2020 doi: 10.1164/rccm.202007-2701OC [published Online First: 2020/10/24]

27. Schulzke SM, Hall GL, Nathan EA, et al. Lung volume and ventilation inhomogeneity in preterm infants at 15-18 months corrected age. J Pediatr 2010;156:542–9 e2.

