## Supplementary material for "Extubation generates lung volume inhomogeneity in preterm infants": Online Supplementary Table 1

63 eligible infants <32 weeks’ GA

30 studied as part of original study

227 eligible infants <32 weeks’ GA

2 excluded (EIT artefact)

12 consent to study at extubation

10 included in analysis

2 excluded (1 unplanned extubation, 1 HFOV)

30 infants included in final dataset

22 consent obtained

20 included in analysis

137 not approached

68 declined consent

Study 1 (Ref #10)

Study 2 (Ref #12)

**Supplementary Figure 1.** Study flow chart
